# LONG-TERM CLINICAL OUTCOMES IN SURVIVORS OF CORONAVIRUS OUTBREAKS AFTER HOSPITALISATION OR ICU ADMISSION: A SYSTEMATIC REVIEW AND META-ANALYSIS OF FOLLOW-UP STUDIES

**DOI:** 10.1101/2020.04.16.20067975

**Authors:** Hassaan Ahmed, Kajal Patel, Darren Greenwood, Stephen Halpin, Penny Lewthwaite, Abayomi Salawu, Lorna Eyre, Andrew Breen, Rory O’Connor, Anthony Jones, Manoj Sivan

## Abstract

**Objective:** To determine the long-term clinical problems in adult survivors of coronavirus (CoV) infection [Coronavirus disease 2019 (COVID-19), Severe Acute Respiratory Syndrome (SARS) and Middle East Respiratory Syndrome (MERS)] after hospitalisation or Intensive Care Unit (ICU) admission.

**Design:** Systematic review and meta-analysis of the literature.

**Data sources:** Ovid MEDLINE, EMBASE, CINAHL Plus and PsycINFO were searched using the strategy: (Coronavirus OR Coronavirus Infections OR COVID OR SARS virus OR Severe acute respiratory syndrome OR MERS OR Middle east respiratory syndrome) AND (Follow-up OR Follow-up studies OR Prevalence). Original studies reporting the clinical outcomes of adult survivors of coronavirus outbreaks two months after discharge or three months after admission were included. The quality of the studies was assessed using the Oxford Centre for Evidence-Based Medicine (OCEBM) 2009 Level of Evidence Tool. Meta-analysis was conducted to derive pooled estimates of prevalence and severity for different outcomes at time points up to 6 months follow-up and beyond 6 months follow-up.

**Results:** The search yielded 1169 studies of which 28 were included in this review. There were 15 Level 1b, 8 Level 2b, 2 Level 3b and 3 Level 4 studies by OCEBM grading. Pooled analysis of studies revealed that complications commonly observed were impaired diffusing capacity for carbon monoxide (DLCO) [prevalence of 27.26%, 95% CI 14.87 to 44.57] and reduced exercise capacity [(6-minute walking distance (6MWD) mean 461m, 95% CI 449.66 to 472.71] at 6 months with limited improvement beyond 6 months. Coronavirus survivors had considerable prevalence of psychological disorders such as post-traumatic stress disorder (PTSD) [38.80%, CI 30.93 to 47.31], depression [33.20%, CI 19.80 to 50.02] and anxiety [30.04%, CI 10.44 to 61.26) beyond 6 months. These complications were accompanied by low Short Form 36 (SF-36) scores at 6 months and beyond indicating reduced quality of life which is present long-term.

**Conclusions:** The long term clinical problems in survivors of CoV infections (SARS and MERS) after hospitalisation or Intensive Care Unit (ICU) admission include respiratory dysfunction, reduced exercise capacity, psychological problems such as PTSD, depression and anxiety, and reduced quality of life. Critical care, rehabilitation and mental health services should anticipate a high prevalence of these problems following COVID-19 and ensure their adequate and timely management with the aim of restoring premorbid quality of life.

## INTRODUCTION

Coronavirus disease 2019 (COVID-19) is the third and largest outbreak of coronavirus (CoV) this century^1^. The disease is caused by severe acute respiratory syndrome coronavirus-2 (SARS-CoV- 2)^2^ with the first cases reported in December 2019^3^. The World Health Organisation (WHO) declared the outbreak as a pandemic on 11th March 2020^4^, with currently more than 2 million infected cases reported worldwide^5^. Infection can lead to severe acute respiratory distress requiring critical care management, and case fatality is around 4%^6^. As a result, much of the current effort is duly focused on improving mortality and ensuring intensive care units and hospital beds are not overwhelmed.

Coronavirus infection results in significant long-term morbidity not only through direct pathology, but also due to secondary disability and iatrogenic complications of treatments^7^. Even though these affect only some survivors^7^, the high prevalence of the disease means it will likely increase healthcare utilization significantly. Therefore, it is necessary to identify the prevalence of these long-term outcomes to facilitate timely preparations for the management of survivors. Whilst few studies are available yet on the long-term outcomes of COVID-19, the two previous CoV outbreaks^1^ of Severe Acute Respiratory Syndrome (SARS) caused by SARS-CoV, originating in Guangdong, China in 2002, and Middle East Respiratory Syndrome (MERS) caused by MERS-CoV, originating in Saudi Arabia in 2012, could be used to model the longer-term impairments of the current pandemic.

This review aims to determine the long-term clinical complications in hospitalised survivors of SARS and MERS. The findings of this review will inform physicians about potential issues prevalent in survivors, help plan appropriate interventions and prepare health and social care services for subsequent increased healthcare utilization post-COVID-19.

## METHODS

### Search Strategy

A search of current literature was carried out in four databases – MEDLINE (1946 to March Week 3 2020), EMBASE (1974 to March 31^st^, 2020), CINAHL Plus (1937 to March Week 3 2020) and PsycINFO (1806 to Match Week 3 2020). The search strategy used was: (Coronavirus OR Coronavirus Infections OR COVID OR SARS virus OR Severe acute respiratory syndrome OR MERS OR Middle east respiratory syndrome) AND (Follow-up OR Follow-up studies OR Prevalence). Terms were entered as MeSH terms where available for each database, otherwise these were searched as keywords in the title, abstract and subject headings.

### Inclusion and Exclusion Criteria

#### Population

Clinical studies involving adults with a confirmed diagnosis of coronavirus infection were included.

#### Exposure

Studies reporting patients with SARS, MERS or COVID-19 from current or previous outbreaks were included.

#### Study Design

Studies had to follow-up patients for a minimum period of 2 months post-discharge or 3 months post-admission to be included in this review. Only primary research studies were included. Reviews, case-reports and editorial reports were excluded.

#### Outcomes

Studies were required to monitor changes in clinical symptoms at follow-up in order to be included. Studies which only monitored changes in serological or immunological results without any assessment of clinical status of the patient were excluded. Likewise, studies reporting only radiological appearance of lung disease or osteonecrosis without any mention of any clinical outcomes were also excluded

### Selection Process

All studies were first screened using the title and abstract. At this stage abstracts with any mention of follow-up were included to avoid exclusion of abstracts which did not report the length of follow-up. Similarly, abstracts which reported follow-up for any outcome were accepted in order to allow inclusion of studies where clinical findings were not significant and therefore not reported in the abstract. Full texts of selected abstracts were then screened to ensure all the above selection criteria were met.

The finalised studies were then critically appraised and graded. Screening and grading were undertaken by four independent reviewers, KP, HA, MS and SH, and DG was involved in cases of disagreement.

### Data Extraction

Data was extracted into standardised tables for each medical system. The following data was extracted: study, year, country, coronavirus outbreak, samples size, follow-up rate, age, sex, settings (hospital/ICU admission), follow-up period, prevalence of key outcomes, mean score for assessment of each outcome. Where estimates were only provided separately for 2 or more subgroups, we took the weighted average across those subgroups as the estimate for the overall population. Extraction was undertaken by at least two independent authors and further cross-checked by two more authors.

### Quality Assessment

Studies were graded using the Oxford Centre for Evidence-Based Medicine (OCEBM) 2009 Level of Evidence Tool [Table 1]^8^. The initial level of evidence was assigned depending on the type of study. Prospective cohort studies were then graded down if follow-up rate was <80%.

**Table 1.** OCEBM Levels of Evidence.

| Level of Evidence | Type of Study |
| --- | --- |
| Level 1a | Systematic review of prospective cohort studies |
| Level 1b | Prospective cohort study with good follow-up (>80%) |
| Level 1c | All or none case-series |
| Level 2a | Systematic review of retrospective cohort study |
| Level 2b | Prospective cohort study with follow-up (<80%) or Retrospective cohort study |
| Level 2c | Ecological studies |
| Level 3a | Systematic Review of Non-consecutive cohort study or very limited population |
| Level 3b | Non-consecutive cohort study |
| Level 4 | Case-series |
| Level 5 | Expert opinion without explicit critical appraisal, or based on physiology, bench research or “first principles” |

### Data Analysis

Binary data for prevalence of outcomes were pooled using meta-analysis by mixed-effects logistic regression. Mean scores for different outcomes were pooled in a meta-analysis using random effects models^9^. Forest plots were stratified by duration of follow-up (up to 6 months and over 6 months). Where a study presented more than one result within a subgroup, we selected the value closest to 6 months (for up to 6 months) or to 12 months (for over 6 months). Between-study heterogeneity was assessed as the range of study estimates, and the proportion of total variability attributable to between-study^10^. There were too few studies to formally explore the sources of heterogeneity through meta-regression (e.g. by mean age, disease, % male, or level of evidence) or examine potential small-study effects such as publication bias through funnel plots. All statistical analyses were conducted using Stata version 15^11^.

## RESULTS

### Study Selection

1169 studies were identified from the databases. Of these 104 abstracts were selected for full-text screening and finally 28 included in the review. The reasons for exclusion of the studies have been reported in Figure 1.

**Figure 1.**
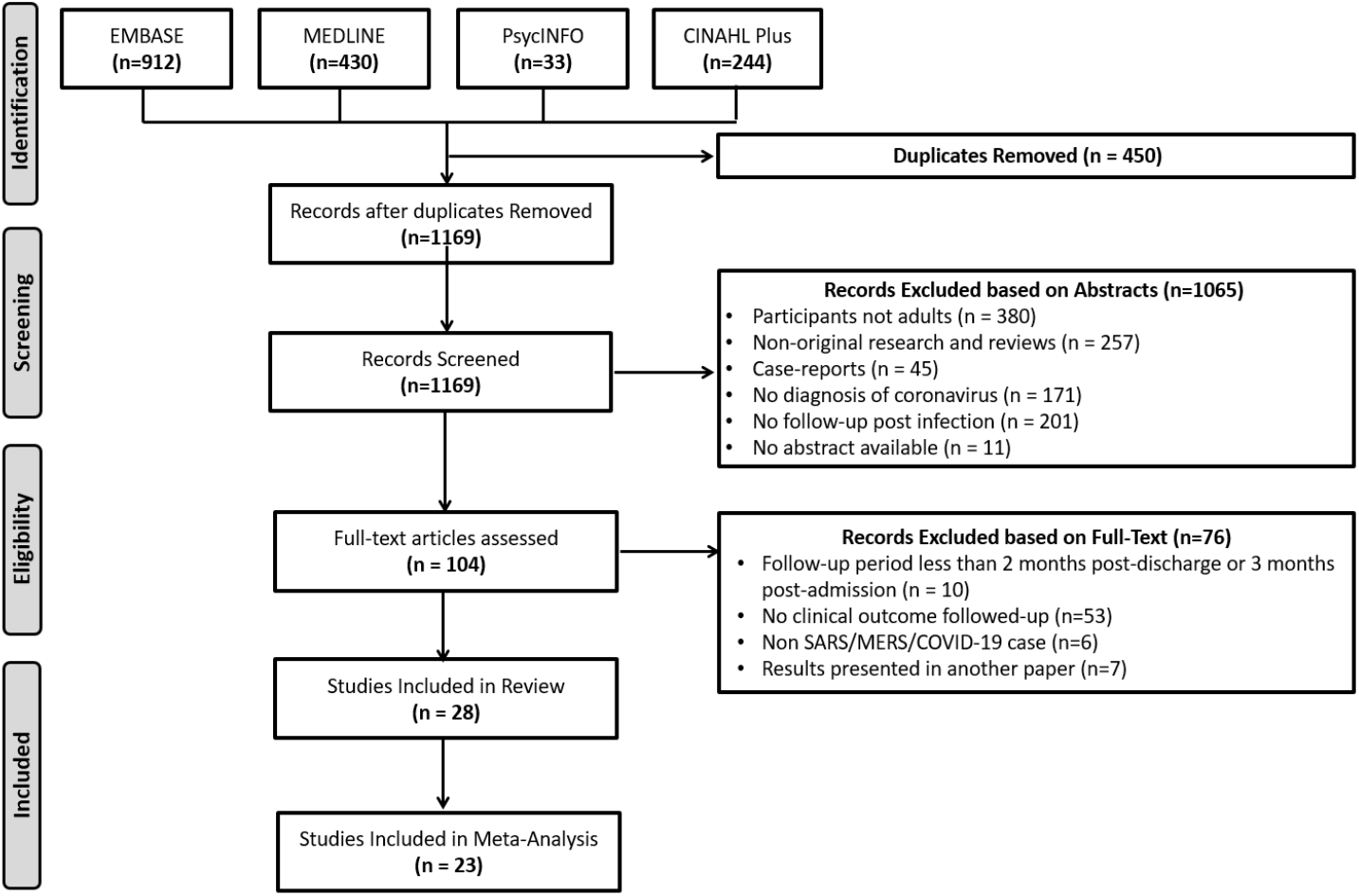
PRISMA Flowchart for the Literature Search.

### Study Characteristics

Out of the 28 studies included in this review, 26 studies reported findings from the SARS outbreak and 2 studies reported findings from the MERS outbreak. No studies have yet reported long-term outcomes of COVID-19 infection. The cohorts studied were from Beijing (11 studies), Hong Kong (9 studies), Guangzhou (1 study), Singapore (2 studies), Taiwan (2 studies), Korea (2 studies) and Canada (1 study) since these were the regions which have been severely affected by the previous outbreaks. The sample size ranged from a case series of 4 patients to a cohort study of 406 patients. There were 15 studies of Level 1b, 8 studies of Level 2b, 2 studies of Level 3b and 3 studies of Level 4 based on OCEBM grading. The 28 studies in the review reported outcomes involving multiple organ systems. The studies mainly addressed one or more of 5 key outcomes of interest – Lung function (18 studies), mental health (6 studies), exercise tolerance (5 studies), health-related quality of life (HRQoL) (5 studies), ocular (1 study) and neuromuscular outcomes (1 study). These are presented in Tables 2-6.

**TABLE 2.**
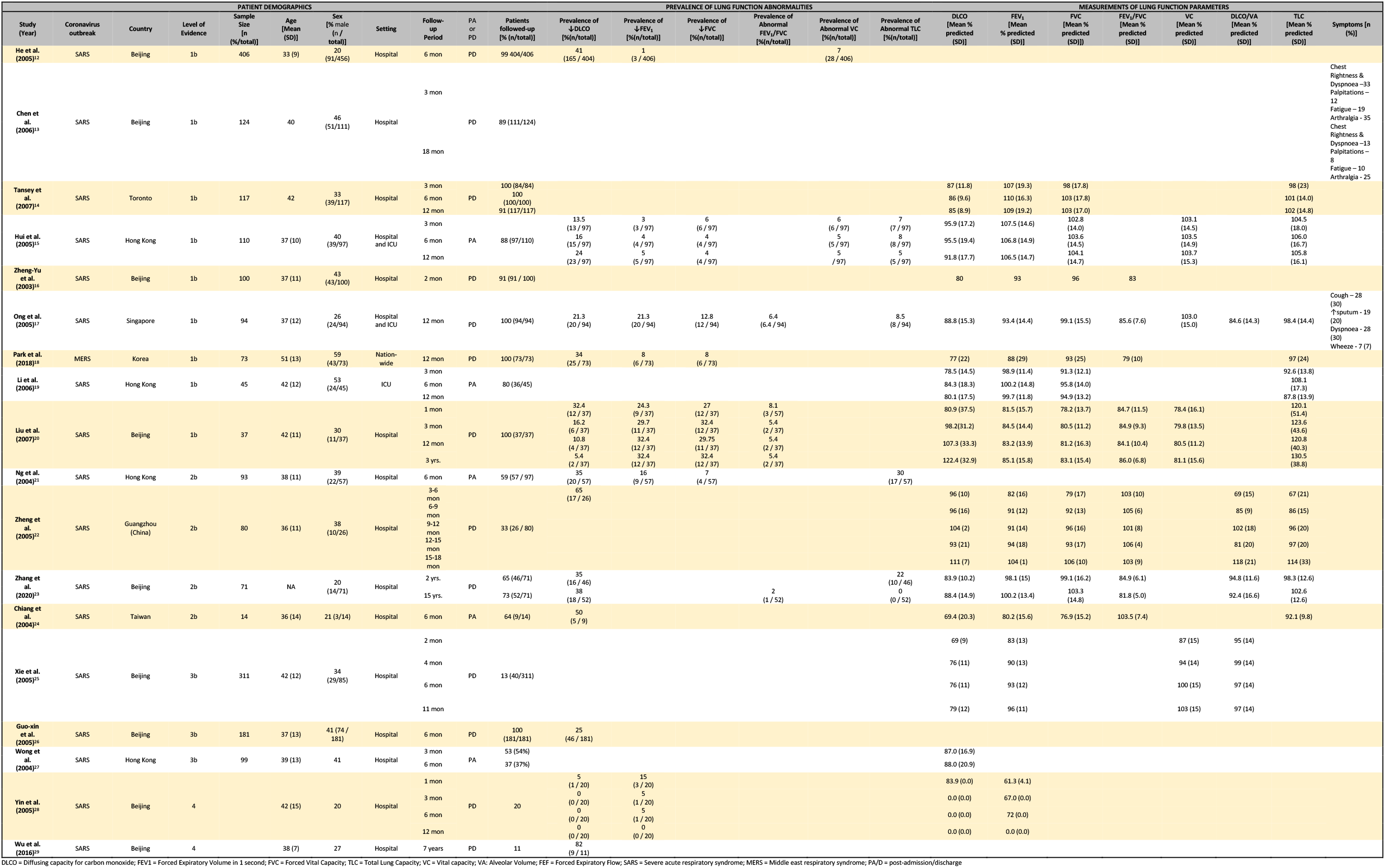
LUNG FUNCTION OUTCOMES.

**TABLE 3.**
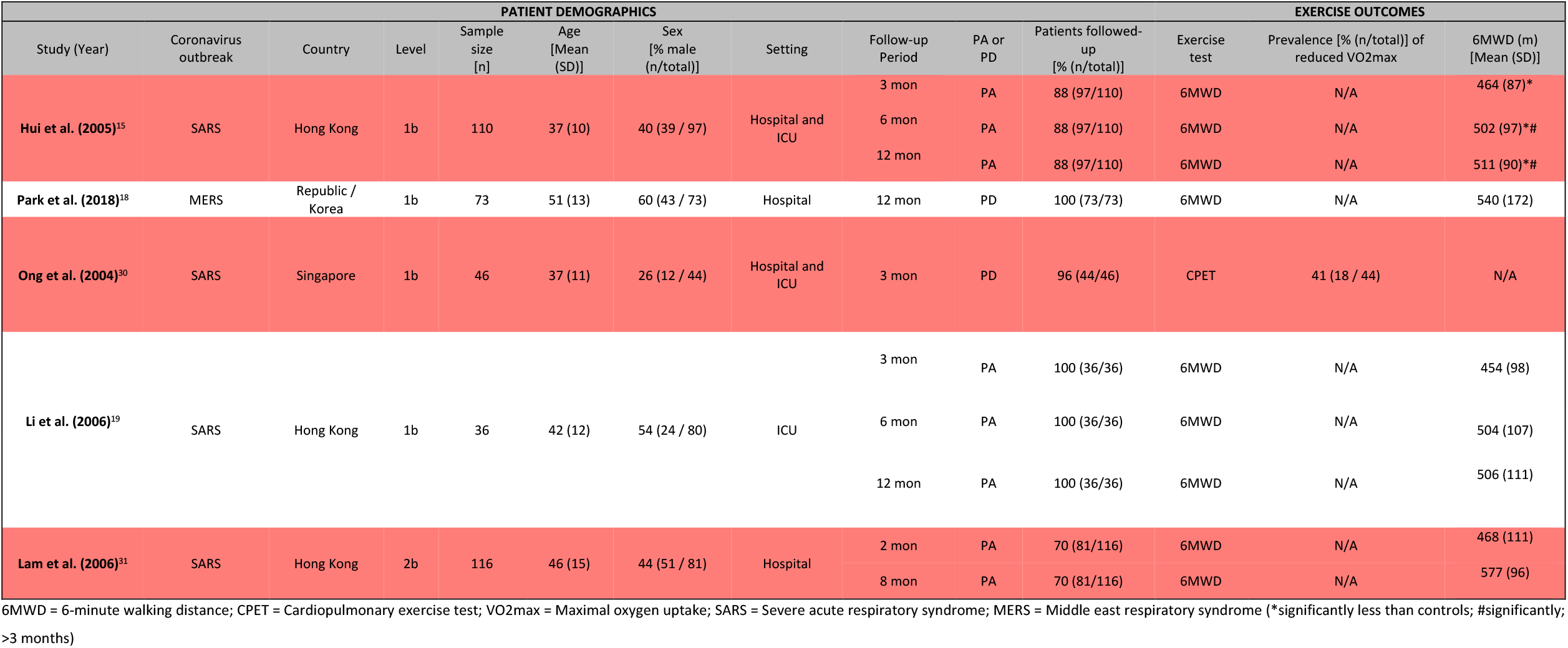
EXERCISE TOLERANCE OUTCOMES.

**TABLE 4.**
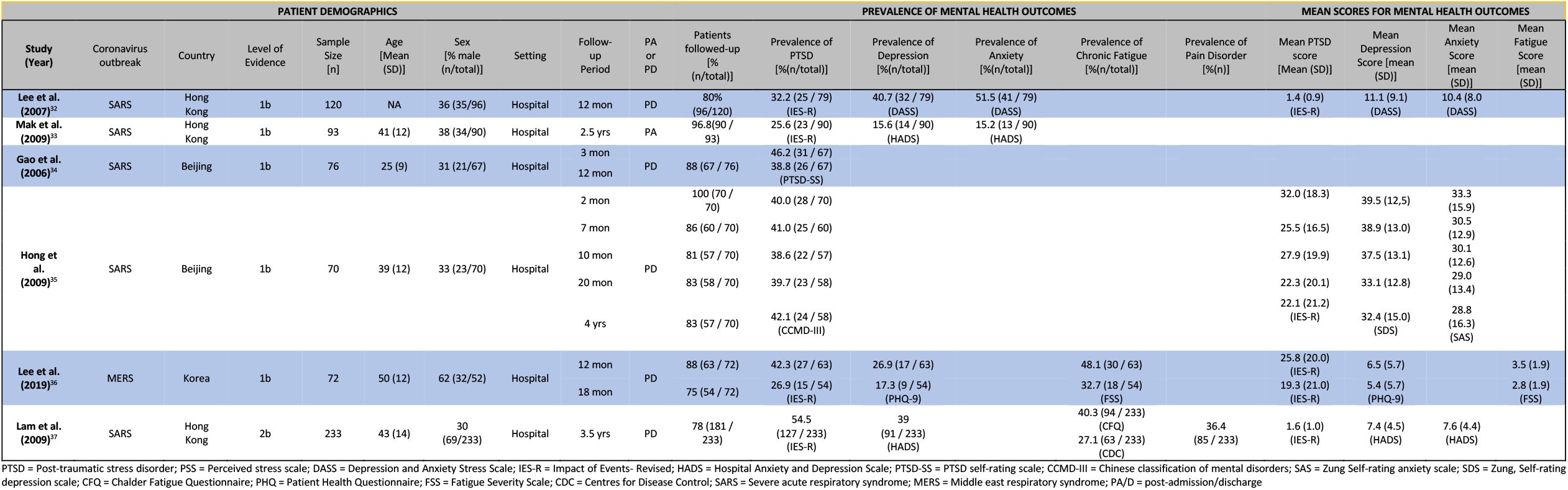
MENTAL HEALTH OUTCOMES.

**TABLE 5.**
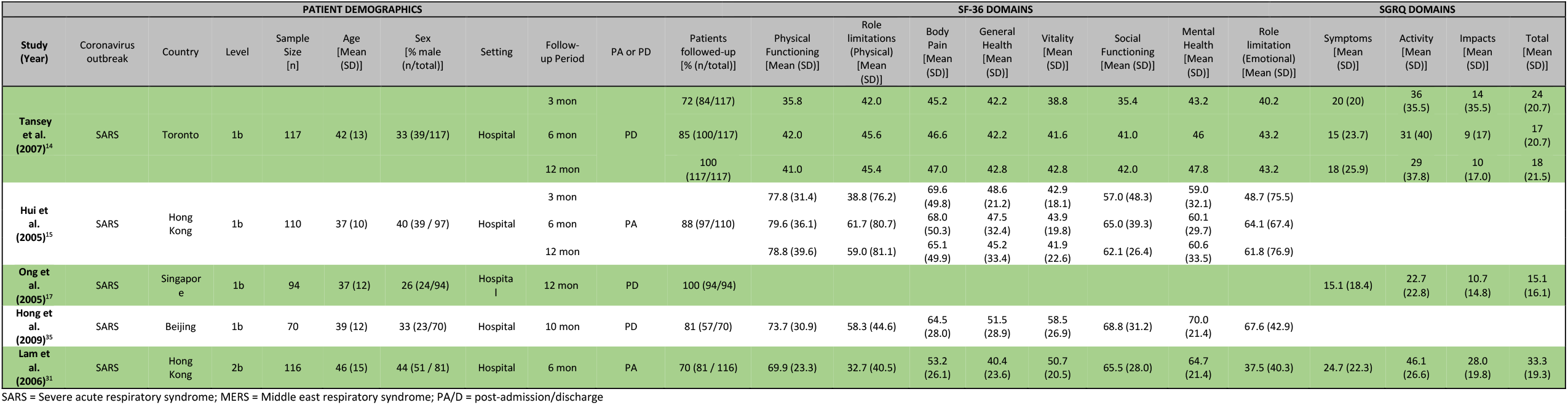
QUALITY OF LIFE OUTCOMES.

**TABLE 6.** MISCELLANEOUS OUTCOMES.

| Study (Year) | Coronavirus Outbreak | System | Country | Level | Sample Size [n] | Age [Mean (SD)] | Sex [% male (n/total)] | Setting | Follow-up Period | Post-admission (PA) or Post-discharge (PD) | Patients followed-up [% (n/total)] | Findings |
| --- | --- | --- | --- | --- | --- | --- | --- | --- | --- | --- | --- | --- |
| Yuen et al. (2004)<sup>39</sup> | SARS | Ophthalmic | Hong Kong | 2b | 45 | 39 (13) | 38 (17/45) | Hospital | 2 mon<br>3 mon | PD | 60 (27/45)<br>33 (15/45) | Elevated intraocular pressure: 2 patients (baseline), 2 patients (2 mon), 1 patient (3 mon)<br>No ocular manifestations of coronavirus were observed |
| Tsai et al. (2004)<sup>40</sup> | SARS | Neuromuscular | Taiwan | 4 | 4 | 46 (7) | 25 (1 / 4) | ICU | 3 mon | PA | 100 (4/4) | Series of patients developed distal-predominant weakness of 4 limbs, mild hyporeflexia and hypesthesia in legs on day 21, 22, 24 and 25 with recovery of muscle power in all patients at 3 months follow-up |

### Lung Function Outcomes

18 studies (9 Level 1b, 4 Level 2b, 3 Level 3b, 2 Level 4 studies) reported lung function outcomes in CoV survivors of which 16 were included in the meta-analysis. Chen et al. (2006)^13^ only reported changes in symptoms without any report of lung function parameters which could be included in this meta-analysis. Zheng-Yu et al. (2003)^16^ did not report standard deviations, hence, the data could not be used in the meta-analysis. Studies reporting prevalence of diffusing capacity of the lung for carbon monoxide (DLCO) (10 studies), forced expiratory volume in 1 second (FEV1) (6 studies), forced vital capacity (FVC) (5 studies) and total lung capacity (TLC) (4 studies) abnormalities were used to pool prevalence of each abnormality [Figure 2 and 3]. At 6 months, abnormalities in DLCO, FVC and TLC were more prevalent than abnormalities in FEV1. Most of these abnormalities improved after 6 months, however, the prevalence of DLCO impairment remained considerably high even 6 months post-infection, with pooled estimate of 24.35 (95% confidence interval 11.05 to 45.46). Studies reporting mean value for DLCO (10 studies), FEV1(10 studies), FVC (10 studies), FEV1/FVC (6 studies), vital capacity (VC) (4 studies), diffusing capacity of the lung for carbon monoxide: Alveolar ventilation (DLCO:Va) (3 studies) and TLC (8 studies) were used to pool mean value for each abnormality up to and beyond 6 months [Figure 4]. The pooled estimates for none of these mean parameters were <80% of predicted.

**Figure 2.**
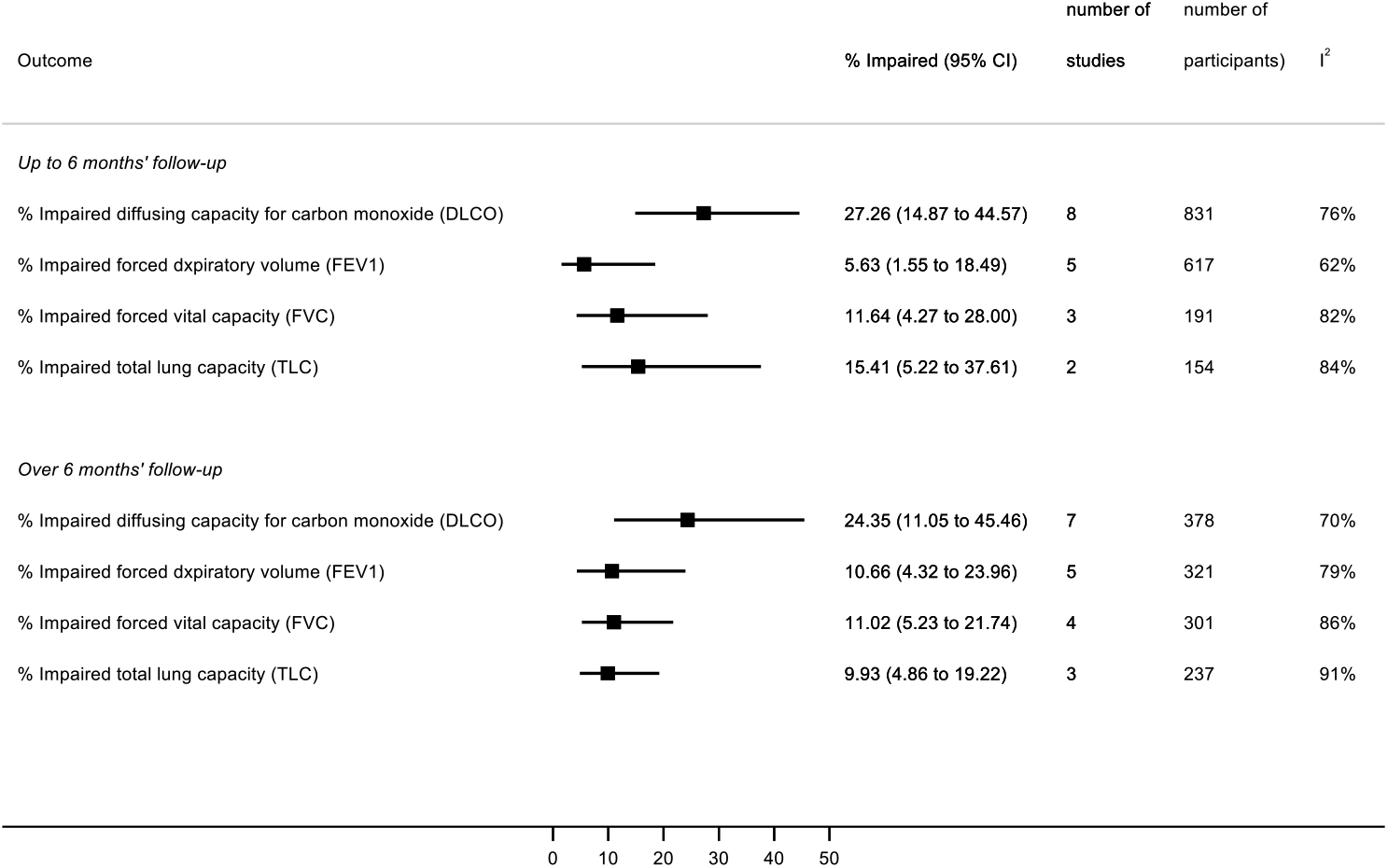
Summary plot showing pooled estimate of prevalence of different lung function abnormalities in CoV survivors up to 6 months (top) and over 6 months (bottom)

**Figure 3.**
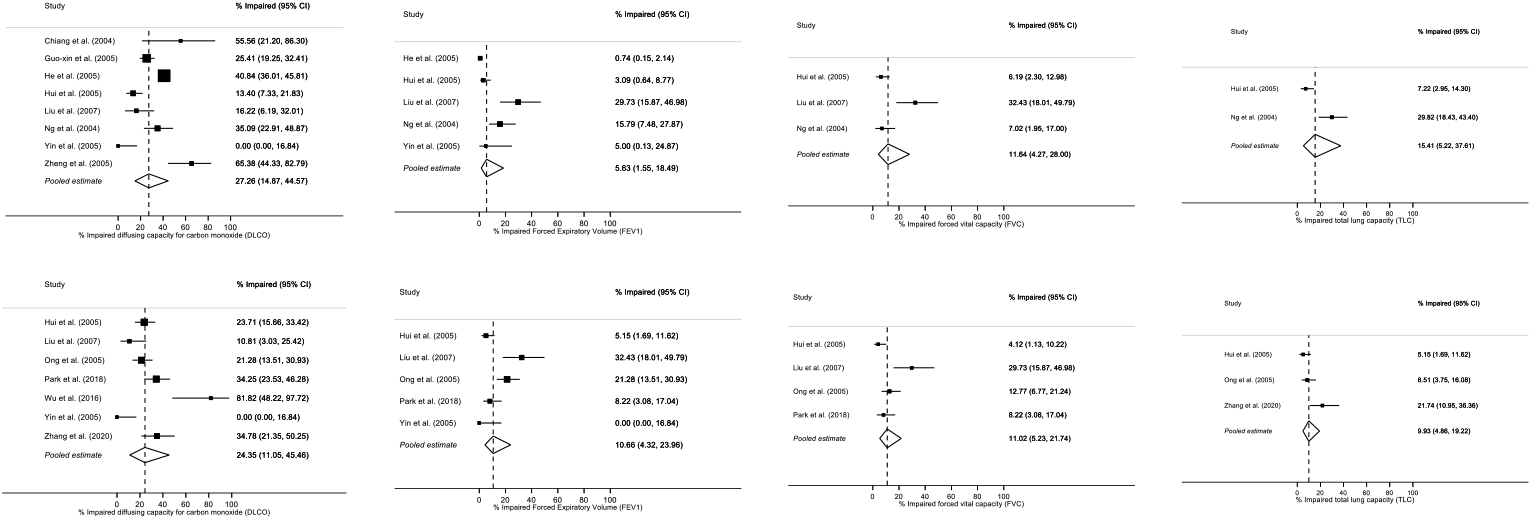
Forest plot showing pooled estimate of prevalence of different lung function abnormalities in CoV survivors up to 6 months (top) and over 6 months (bottom)

**Figure 4.**
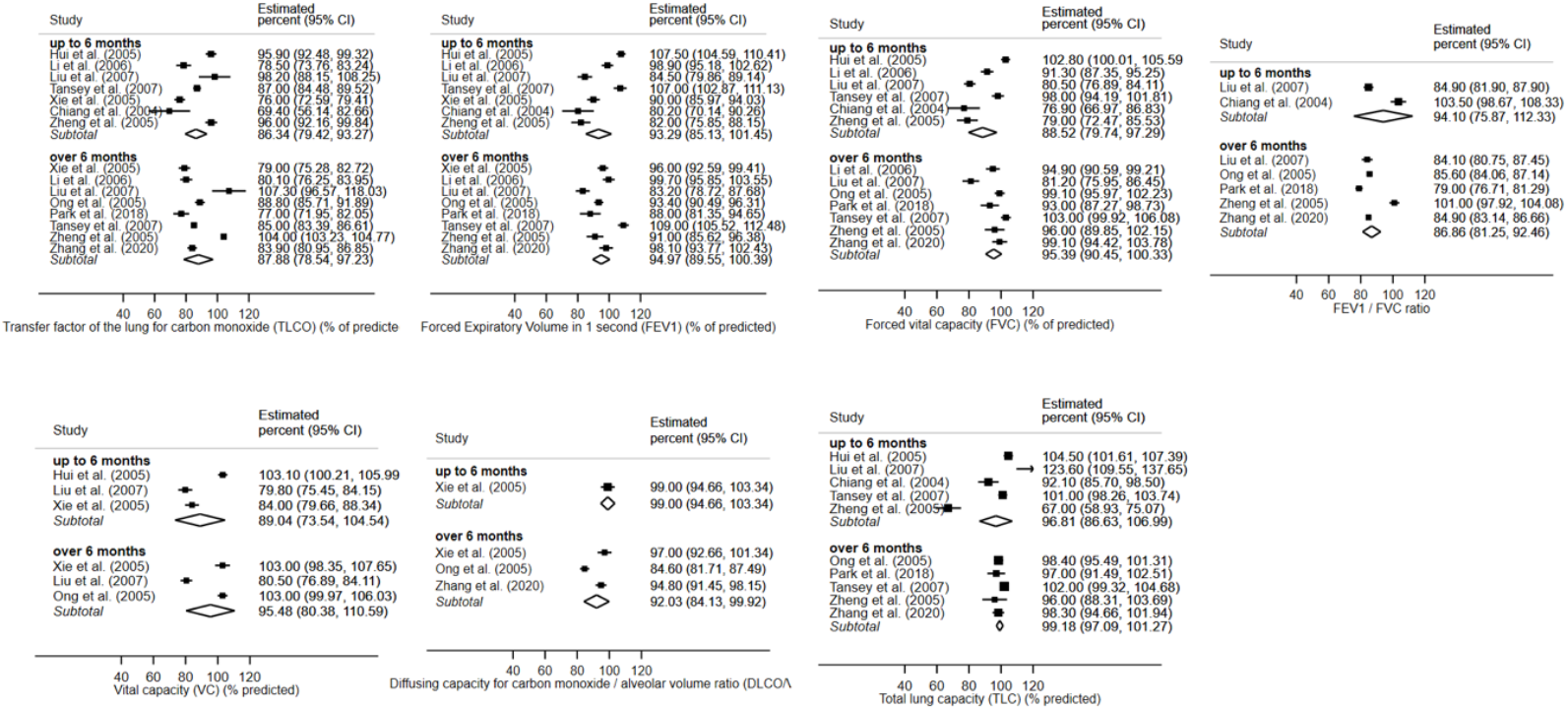
Forest plot showing pooled estimate of mean values of different lung function abnormalities in CoV survivors up to 6 months (top) and over 6 months (bottom)

### Exercise Tolerance Outcomes

5 studies (4 Level 1b studies and 1 Level 2b study) reported exercise tolerance outcomes in CoV survivors of which 4 were included in this meta-analysis [Figure 5]. Results from Ong et al. (2004)^30^ were not included because they only reported outcomes from cardiopulmonary exercise testing (CPET) and did not conduct 6-minute walking distance (6MWD). The pooled estimate of 6MWD for 3 studies reporting outcomes up to 6 months was 461.18 (95% Confidence Interval 449.66 to 472.71). The 6MWD increased substantially after 6 months with pooled estimate of 533.00 (95% Confidence Interval 449.66 to 472.71). Since ∼30 m is considered to be the minimal clinically important difference in 6MWD^41^, patients seem to improve significantly overtime. Unfortunately, data was not available regarding the 6MWD for participants before CoV infection and therefore there is no report of the number of patients with exercise tolerance lower than baseline.

**Figure 5.**
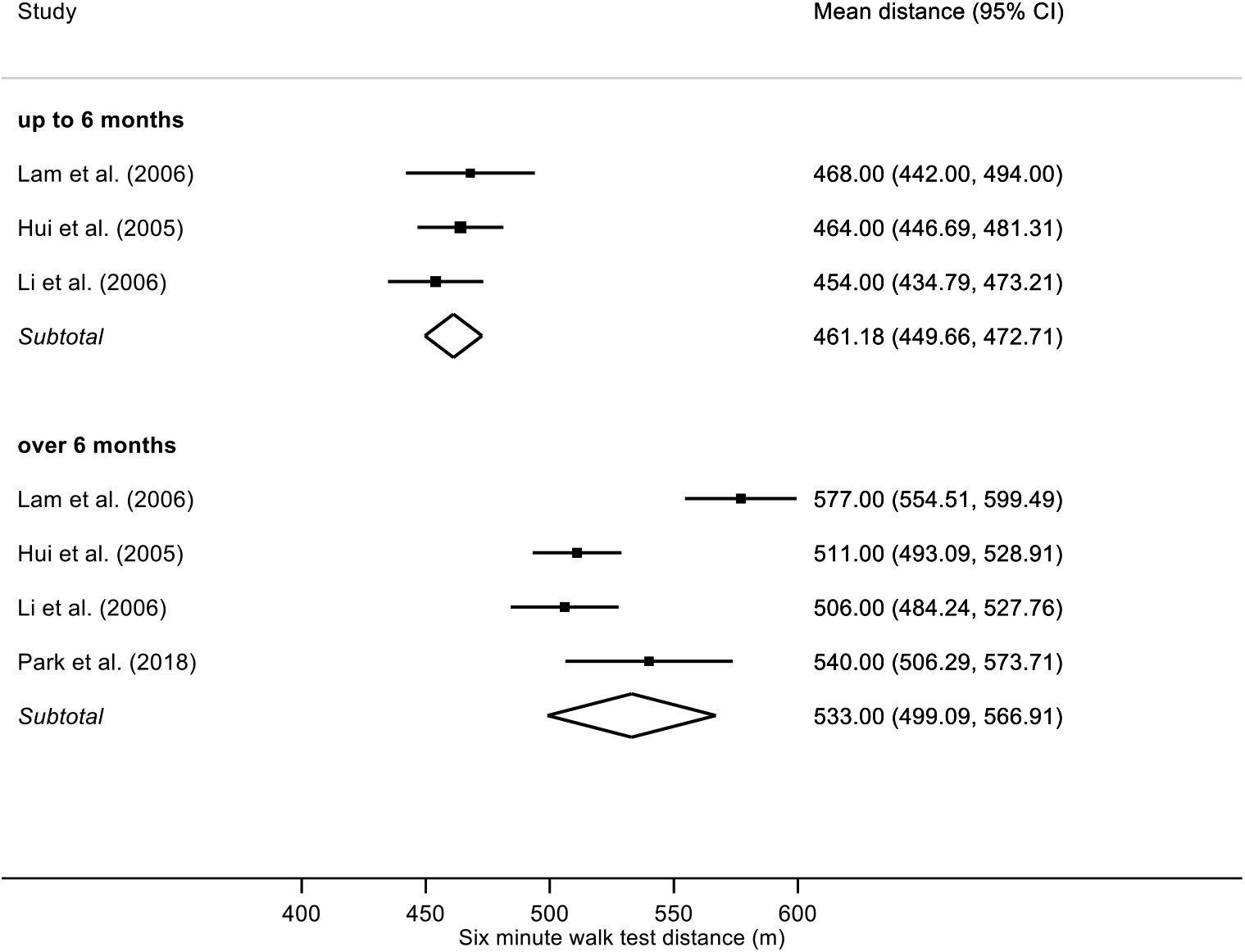
Summary plot showing pooled estimate of 6-minute walking distance in CoV survivors up to 6 months (top) and over 6 months (bottom)

### Mental Health Outcomes

5 studies (5 Level 1b and 1 Level studies) reported psychological comorbidities in CoV survivors of which all 6 were included in the meta-analysis. All studies which reported prevalence of these psychological conditions had follow-up period of longer than 6 months. As a result, meta-analysis was conducted for prevalence beyond 6 months only [Figure 6 and 7]. The prevalence of different psychological conditions was substantially high with pooled estimates of 38.80% (95% confidence interval 30.93 to 47.31) for post-traumatic stress disorder (PTSD), 33.20% (95% confidence interval 19.80 to 50.05) for depression and 30.04% (95% confidence interval of 10.44 to 61.26) for anxiety [Figure 6 and Figure 7]. We could not perform meta-analysis on the mean scores for different psychological comorbidities because different scales were used by different studies to report these.

**Figure 6.**
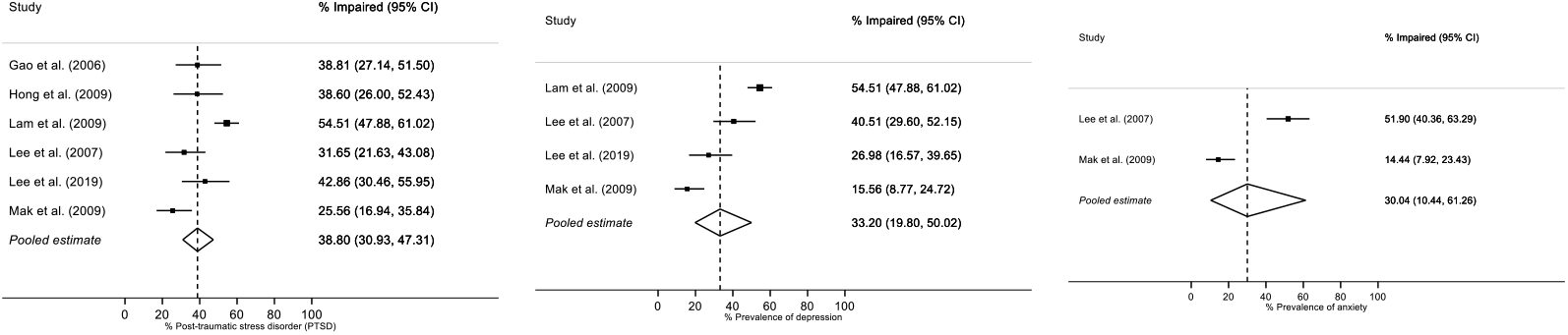
Forest plot showing pooled estimate of prevalence of different psychological conditions in CoV survivors over 6 months.

**Figure 7.**
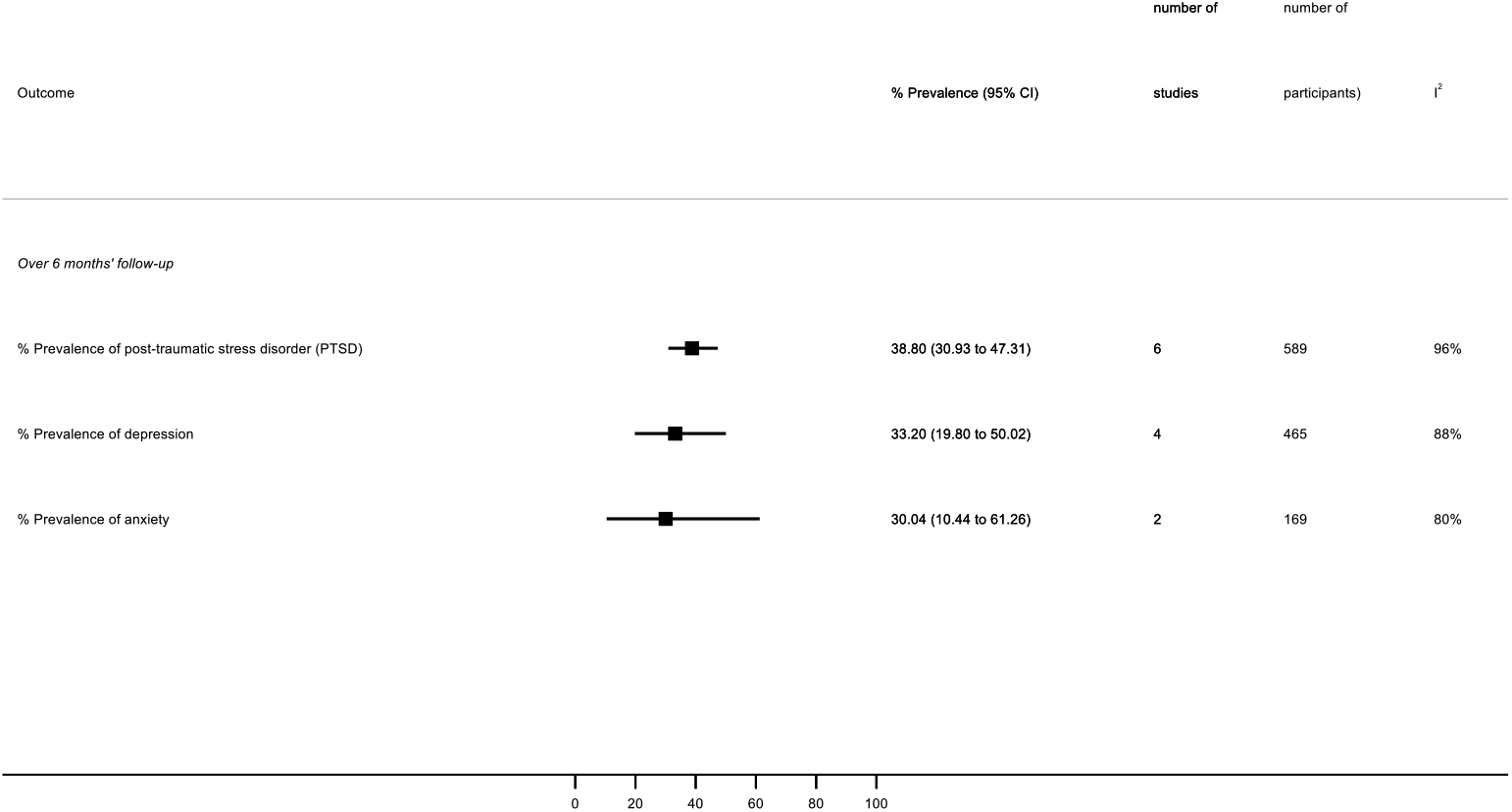
Summary plot showing pooled estimate of prevalence of different psychological conditions in CoV survivors over 6 months.

### Quality of Life Outcomes

5 studies (4 Level 1b studies and 1 Level 2b study) reported quality of life outcomes in CoV survivors. Out of these, only 3 studies, which reported both mean and SD, were included in the meta-analysis of short form 36 health survey (SF-36) [Figure 8 and 9] and St George’s Respiratory Questionnaire (SGRQ) [Figure 10] each. The pooled analysis showed that the mean score for all of the 8 domains of the SF- 36 were substantially lower in CoV survivors than normative values for people who are healthy as well as for people with chronic diseases derived from existing validated literature^42^[Figure 9]. Domains which scored particularly lower than healthy individuals and those chronic conditions were role limitations due to physical and emotional health. There seems to be some improvement in these domains beyond 6 months, but the scores were still lower than healthy and chronic disease patients^42^.

**Figure 8.**
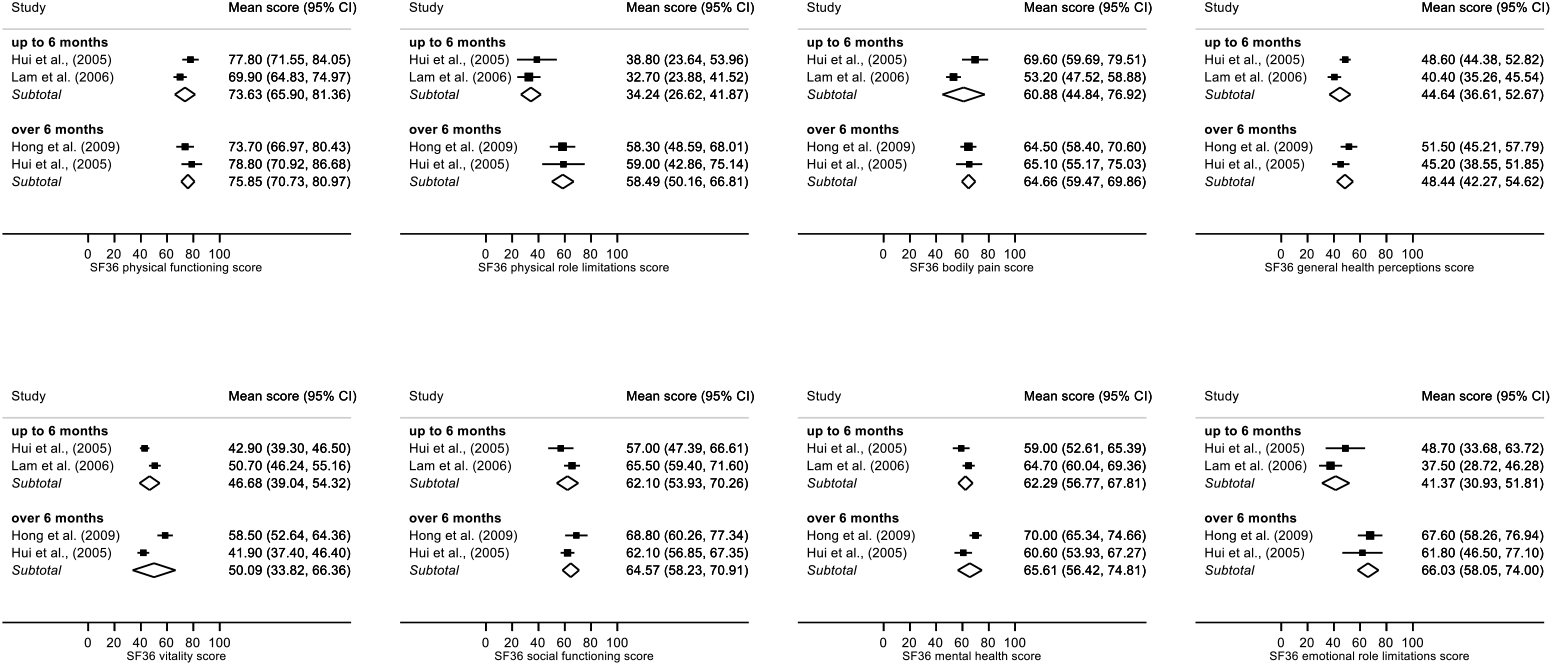
Forest plot showing pooled estimate of mean score for different domains of SF-36 in CoV survivors up to 6 months (top) and over 6 months (bottom)

**Figure 9.**
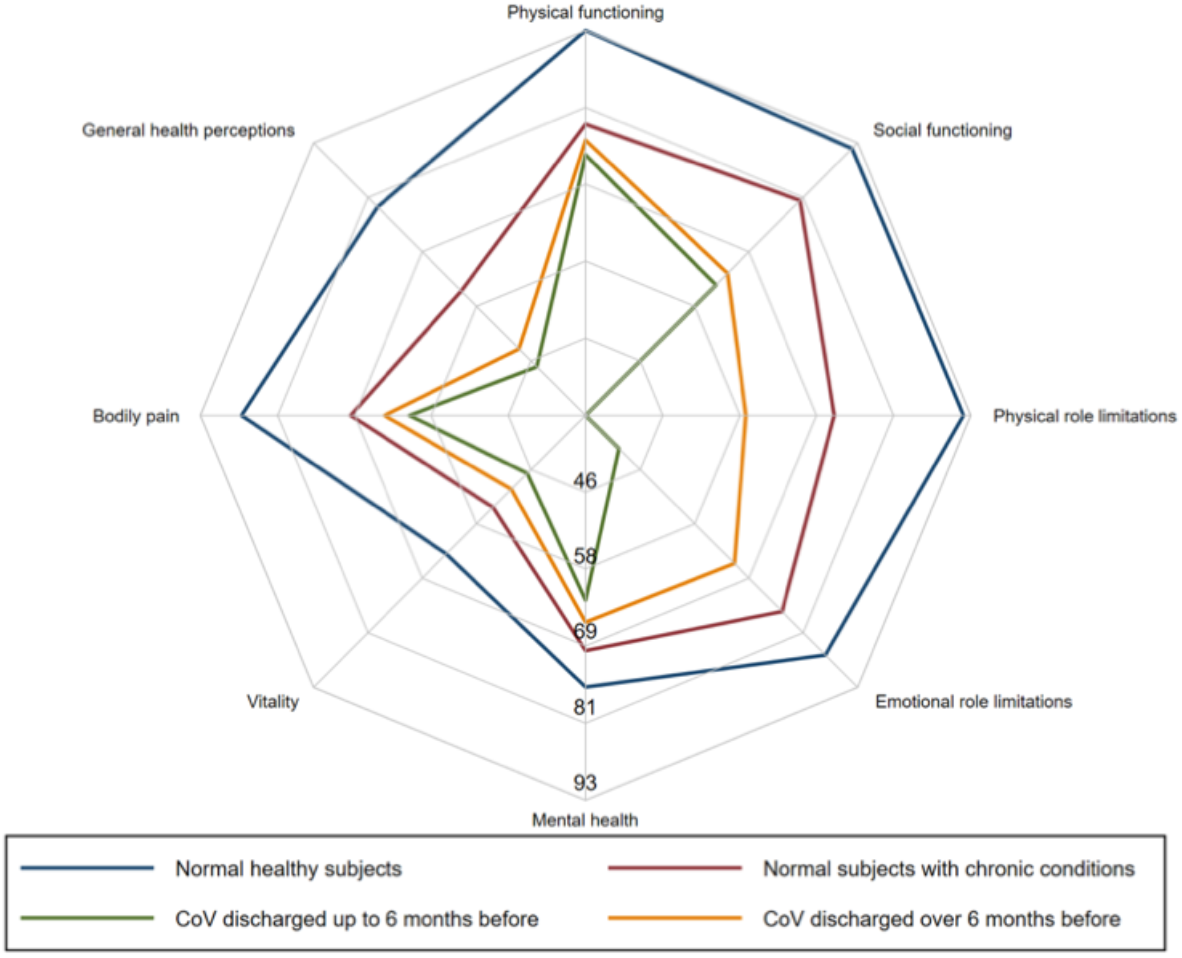
Radar plot showing pooled estimate of mean scores for different domains of SF-36 in CoV survivors up to 6 months (green) and over 6 months (orange) compared to healthy individuals (blue) and subjects with chronic conditions (red).

**Figure 10.**
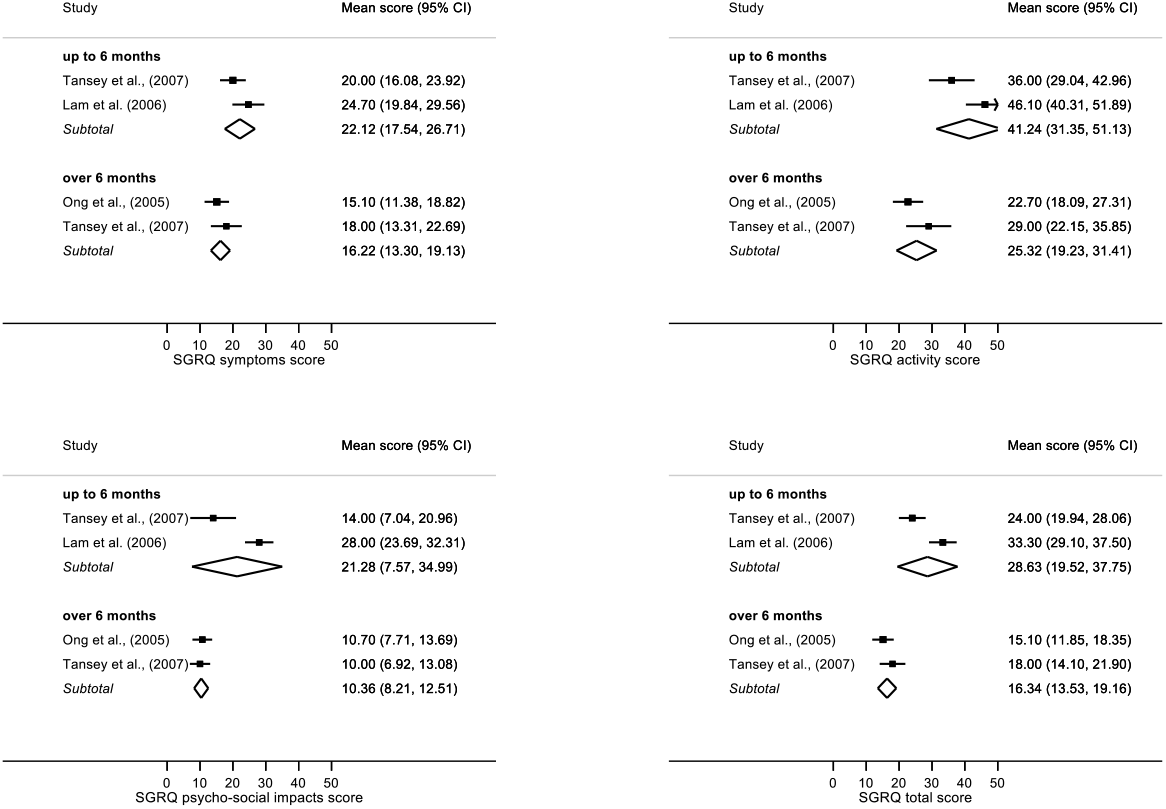
Forest plot showing pooled estimate of mean score for different domains of SGRQ in CoV survivors up to 6 months (top) and over 6 months (bottom)

### Other Outcomes

Other outcomes which have been followed-up in SARS patients have been reported in Table 6. The study by Yuen et al. (2004)^39^ found no eye pathologies in this cohort. The case-series by Tsai et al. (2004)^40^ followed up patients who developed limb weakness related to critical illness neuropathy (CIN) and myopathy (CIM) and sensory deficits following infection. All these patients had a partial or full recovery of muscle power 3 months after admission.

## DISCUSSION

The long-term complications of coronavirus infection are not well understood. The prevalence, severity and prognosis of these complications must be determined to plan the rehabilitation of survivors of the current COVID-19 pandemic. This systematic review collates these long-term complications seen following previous coronavirus outbreaks (SARS and MERS) in those who required hospitalisation or ICU stay. Our findings highlighted that the health-related quality of life (HRQoL), measured using SF-36, is considerably reduced in CoV survivors at 6 months post-infection, shows only slight improvement beyond 6 months and remains below normal population and those with chronic conditions [Figure 9]. As these SF-36 scores reflect impairment in physical, mental and social functioning of well-being, it is not surprising that the key areas of impairments identified in our systematic review were pulmonary dysfunction, reduced exercise tolerance and psychological problems.

Respiratory compromise is one of the key physical issues in survivors. The impairment is mainly restrictive in nature with predominance of abnormalities in DLCO, VC and TLC compared to FEV1, thereby, supporting the etiopathology of acute respiratory distress syndrome with parenchymal infiltration caused by the infection. Even though lung function improves over time, the results from our meta-analysis showed that reduction in DLCO may still be present in 11 to 45% of CoV survivors at 12 months. This is consistent with CT findings of other studies which have reported that pulmonary fibrosis can persist up to 7 years^29^. Pulmonary rehabilitation has been shown to improve QoL in other patients with fibrosis^43^ but it is unknown whether this would be effective in COVID-19 survivors.

CoV survivors had reduced aerobic capacity with peak oxygen uptake (VO2max) testing showing impairments in 41% of patients at 3 months^30^. This could be due to circulatory limitation, muscle weakness, critical illness neuropathy and myopathy (CINM) and deconditioning^30^. The 6MWD is also reduced at 3 months and slowly improves by 12 months^44,45^. We know from other literature that such chronic weakness may be present in patients even 5 years after ICU admission, therefore, rehabilitation needs of these patients can be prolonged^46^. Early rehabilitation combining mobilisation with strengthening exercises may improve exercise tolerance in these patient groups as it has substantial evidence for improving weakness and functional independence in CINM^47^.

Our meta-analysis showed that around a third of CoV survivors may have psychological conditions such as PTSD, depression and anxiety beyond 6 months. These estimates are much higher than the prevalence of these conditions reported as part of post-ICU syndrome in medical and surgical patients^48^. This indicates that the long-lasting mental health impact is not from serious illness alone, but also from factors such as fear^49^, stigma^37^ and quarantine^50^, all of which also apply to COVID-19^51^. The neuropsychiatric aspects of CoV infections are not very well known yet and priorities and strategies for mental health science research have already been set out^52^.

SF-36 scores for role limitations in CoV survivors were particularly low compared to healthy individuals. Tansey et al. (2007)^14^ reported that 17% CoV survivors had not returned to their previous level of working even at 1 year post-infection. Many of the symptoms experienced by CoV survivors could be responsible for such reduced social functioning. Fatigue was reported to be present in at least a third of the patients in two studies with a follow-up period of 18 months^36^ and 40 months^37^ each. Pain disorders were followed-up in one study which reported it to be present in about one-third of patients^37^.

The main strength of this study is that it highlights multiple long-term biopsychosocial impairments which may hinder return to pre-infection functional status. This is the first systematic review and meta-analysis on this topic as far as we are aware. Unlike a previous review from 2003^7^, we investigated long-term outcomes from major SARS and MERS outbreaks this century. We have tried to capture the various aspects of well-being and health-related quality of life in CoV survivors. There are understandably no studies on the long-term effects of COVID-19 as the outbreak was first reported only in Dec 2019. Considering SARS-CoV-2 belongs to the same virus family and has led to a more rapid spread with greater mortality worldwide^1^, the aftereffects are predicted to be similar, if not more profound and demanding of healthcare resources long term. For example, the widely reported prevalence of coagulopathy and thrombotic disease in COVID-19 patients may result in new end-organ complications and respiratory recovery could conceivably be affected^53^, thereby, leading to worse outcomes long-term.

Finally, there was also a paucity of information following up SARS and MERS survivors. Many studies had a small sample size and some outcomes could not be quantified because of limited number of studies reporting these. There was substantial heterogeneity, with almost all I-squared estimates >50%. We were unable to formally explore sources of this heterogeneity because of the small numbers, but these could include study-level differences in mean ages, gender, differences between SARS and MERS outbreaks, referral pathways between regions and study design.

Differences in outcomes between ICU and non-ICU patients remain unclear. Whilst one study identified that lung function parameters like FVC and DLCO were comparatively lower in ICU group^15^, another reported no significant difference between the two groups^30^. Further reporting of outcomes in ICU CoV survivors would be crucial as muscle weakness developed during ICU admissions has been associated with substantial impairments in physical function and quality of life^54^. Therefore, coronavirus survivors who required ICU will likely have even worse outcomes.

The global community of rehabilitation and mental healthcare services need to address the long-term complications identified in this review very early in the COVID-19 pandemic recovery phase. Acute rehabilitation during hospital stay requires active involvement of multidisciplinary teams to ensure the physical, psychological and social aspects are met. Post-acute early rehabilitation in the first 3 months after discharge is critical to prevent emerging issues such as reduced exercise tolerance and depression. Long-term rehabilitation must be an ongoing process to ensure individual function and biopsychosocial profiles are restored as much as possible so these individuals can return to previous societal roles and start contributing successfully to economies. This will determine whether the healthcare services around the globe have successfully managed the long-term impact of this pandemic.

## Data Availability

All data related to this systematic review and meta-analysis will be made available for audit and cross-checking

